# Global thickness for accurate and precise evaluation of post-intervention remodelling in left ventricular hypertrophy

**DOI:** 10.64898/2026.11.01.26367004

**Authors:** Lynn J Khor, Hunain Shiwani, George D Thornton, Jonathan Bennett, Nikoo Aziminia, Abhishek Shetye, Constantin-Cristian Topriceanu, Gabriella Captur, Rhodri H Davies, James C Moon, Peter Swoboda, Calvin WL Chin, Thomas A Treibel, Rebecca Kozor, Martin Ugander, PRECISION-HCM Collaborative

## Abstract

**Background:** Cardiovascular interventions may impact both left ventricular mass (LVM) and end-diastolic volume (EDV), but the net effect on wall thickness has been difficult to accurately evaluate. Global average left ventricular end-diastolic wall thickness (GT) relates non-linearly to LVM and EDV. We hypothesized that if GT is independent of EDV in health, then GT will be an ideal measure of post-intervention change in left ventricular hypertrophy.

**Methods:** GT [mm] was calculated using LVM and EDV by cardiovascular magnetic resonance as previously validated. The correlation between GT and EDV was evaluated in healthy volunteers. Change in GT following intervention was evaluated for aortic stenosis undergoing valve replacement, hypertension commencing antihypertensive medications, athletes undergoing forced detraining, and aortic regurgitation undergoing valve replacement. The performance of GT was compared to LVM/EDV.

**Results:** Among healthy volunteers (n=4461, mean±SD age 59±10 years, 50% female), EDV correlated with LVM/EDV but negligibly with GT (R^2^>0.16 vs R^2^<0.02, p<0.001 for both sexes). GT and not LVM/EDV changed after intervention in all conditions (p≤0.02 for all). Post-intervention changes in GT varied by condition (mean±SEM aortic stenosis [n=102]: -1.75±0.13mm, hypertension [n=78]: -0.66±0.08mm, athletes [n=28]: -0.29±0.09mm, aortic regurgitation [n=58]: +0.50±0.11 mm, p≤0.02 for all comparisons).

**Conclusions:** In health, GT, unlike LVM/EDV, remains effectively unchanged across EDV values, indicating that GT is a physiological constant in health and can track pathological changes. After intervention, only GT and not LVM/EDV consistently changed. Instead of LVM/EDV, GT should be used to quantify remodelling after interventions in left ventricular hypertrophy.

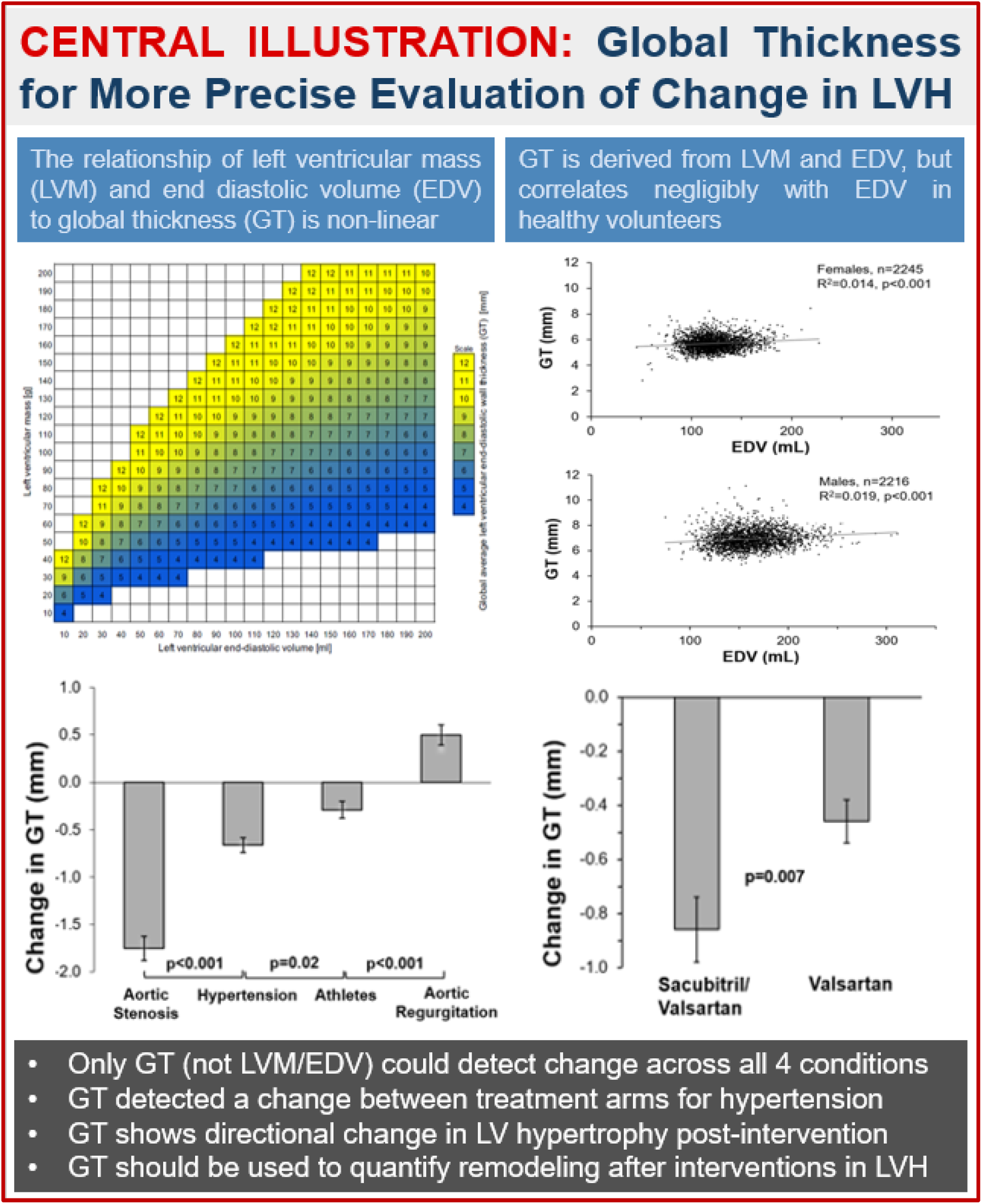

## Introduction

Left ventricular hypertrophy (LVH) is a well-established indicator of poor cardiovascular prognosis [1]. Reverse remodelling in LVH is achievable with various treatments and is associated with improved cardiac function and reduced cardiovascular risk [2]. Consequently, LVH regression has become a common outcome measure in interventional studies involving cardiovascular conditions that cause LVH, such as hypertension [3] [4], infiltrative cardiomyopathies [5] and valvular heart disease [6].

Given its high accuracy and reproducibility, cardiovascular magnetic resonance (CMR) imaging is often regarded as the non-invasive reference standard in the assessment of left ventricular mass and volumes when assessing LVH [7]. LV geometry may vary as changes in left ventricular mass (LVM) affect left ventricular end-diastolic volume (EDV) in a non-linear manner. Therefore, a reduction in overall LVM may reflect a reduction in EDV, and does not always reflect reduction in left ventricular wall thickness.

Commonly used CMR measures of LVH include left ventricular mass index (LVMI), maximum end-diastolic wall thickness, and the ratio between left ventricular mass and left ventricular end-diastolic volume (LVM/EDV, also referred to as mass-to-volume ratio (MVR)). These measures are frequently used to assess the impact of interventions on LVH. However, the non-linear relationship between LVM, EDV and LV wall thickness makes it challenging to accurately detect the net effect on change in LVH over time.

Global average left ventricular end-diastolic wall thickness (GT) has been shown to be accurately and precisely estimated using LVM and EDV [8]. Notably, GT has a non-linear relationship to LVM and EDV (Figure 1). GT estimates the average wall thickness of the whole left ventricle in millimetres (mm) and has been shown to associate with prognosis [8]. LVMI by CMR has been shown to be the strongest prognostic measurement in LVH that relates to death and heart failure hospitalisation, followed by GT. However, in the absence of an increased LVMI, GT is the most prognostic measurement, highlighting its utility in evaluating LVH among subjects with an LVMI within the normal range [8].

**Figure 1.**
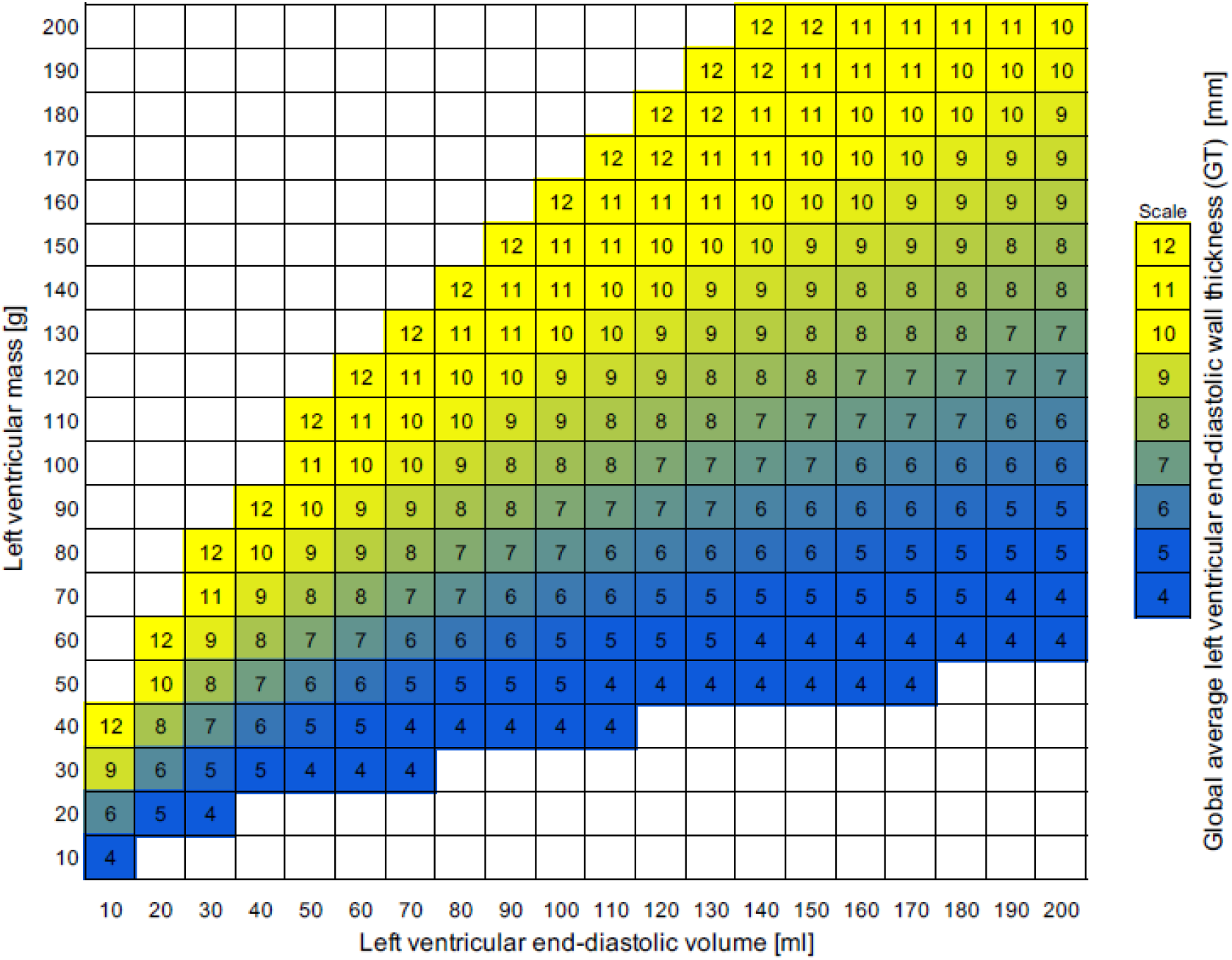
The relationship between left ventricular mass (LVM), left ventricular end-diastolic volume (EDV) and global average left ventricular end-diastolic wall thickness (GT). Note that for a given GT of the same color, there is a non-linear relationship between LVM and EDV. GT is calculated from LVM and EDV as previously empirically validated [8].

In this study, we first explore the relationship between EDV and both GT and LVM/EDV, respectively, in a healthy population. We hypothesize that GT would be unchanged by EDV in health. Next, we sought to determine if interventions in severe aortic stenosis, hypertension, athletes, and severe aortic regurgitation, respectively, lead to changes in GT. We further hypothesized that GT would more reliably detect reverse remodelling in LVH compared to LVM/EDV.

## Methods

### Study population

This is a retrospective study of deidentified CMR data from previously published cohorts. CMR data was obtained from previously studied healthy normal participants with no known cardiovascular diseases [9]. Also, a randomly selected subgroup (n=50) of these subjects was used to evaluate the differences in correlation between GT by CMR and wall thickness estimated from EDV and LVM using different exponents for LVM^x^/EDV^y^ (x and y over the range of 0.01 to 1.00 at intervals of 0.01). This enables comparison with the formulas for GT by CT [10], Concentricity^0.67^ and LVM/EDV.

Next, we analysed CMR data pooled from four different studies of interventions in LVH. Cohorts were chosen from a spectrum of cardiac conditions that are expected to have different underlying mechanisms that lead to LVH. The four cardiac conditions selected include data from previously published studies on severe aortic stenosis treated with aortic valve replacement [11], patients with essential hypertension being treated with antihypertensive medications (either sacubitril/valsartan or valsartan alone) [12], athletes who underwent forced detraining due to incidental limb bone fracture [13], and severe aortic regurgitation treated by aortic valve replacement [14]. All participants were recruited prospectively and underwent CMR before and after intervention as per the respective study protocols. Ethics approvals and written informed consent were obtained at the respective sites. This study was granted exemption from approval by the University of Sydney human subject research ethics office.

### CMR

All five study cohorts used standard CMR clinical scan protocols. CMR measurements of left ventricular volumes and mass were performed by the respective investigator groups. Deidentified CMR measurements of EDV, left ventricular end-systolic volume (ESV), LVM, height, weight, age, and sex were collected. GT and LVM/EDV were computed from deidentified CMR measurements provided by the respective studies. All studies used measurement of left ventricular mass and volumes that used rounded contours that excluded papillary muscles from the LVM.

### Definitions

GT was calculated for all participants using the formula: GT [mm] = 0.05 + 1.60 * LVM[g]^0.84 * EDV [ml]^−0.49 as previously derived and validated [8]. LVM/EDV was calculated as the ratio of LVM to EDV [g/mL]. For selected comparisons, the previously described concentricity^0.67^ measure was calculated as LVM/EDV^2/3^ [15].

### Study outcomes

Differences between LVM, EDV, LVM/EDV and GT before and after intervention were calculated for all four intervention groups. Furthermore, to explore the potential utility of GT in detecting regression of LVH between two different treatment arms, we analysed data separately based on treatment of hypertension with valsartan alone, or the combination of sacubitril and valsartan [12].

### Statistical analysis

Continuous variables were assessed for normal distribution by visual assessment of histograms and Q-Q plots. Data is presented as mean±SD for baseline characteristics of the healthy population and interventional studies, and mean±SEM to illustrate point estimates for effect magnitudes. The paired t-test was used to compare measures pre and post intervention. One-way ANOVA with Games-Howell post-hoc adjustment was used to compare post-intervention differences between populations. The independent samples t-test was used to determine if post-intervention changes differed between treatment arms. Differences in strength of correlation were tested with Steiger’s Z-test. [16] All other statistical analyses were performed using Jamovi version 2.7 (The jamovi project, 2025). A p<0.05 was considered statistically significant.

## Results

### Healthy population

Baseline characteristics of the healthy population are summarized in Table 1. Normal values for GT in the healthy population were 5.71±0.55 in females and 6.95±0.74 in males. LVM/EDV was correlated with EDV for both males and females (R^2^>0.16, p<0.001 for both, Figure 2). By comparison, GT had a negligible correlation with EDV for both males and females (R^2^<0.02, p<0.001 for both, Figure 2). These differences in strength of correlation with EDV were greater for LVM/EDV than GT (p<0.001 for both sexes). When exploring the relative strengths of correlation between GT by CMR and an exhaustive combination of different LVM and EDV exponents, the closest correlations to the CMR GT formula were found for the formula for GT by computed tomography (R^2^=0.998, p<0.001) and concentricity^0.67^ (R^2^ =0.992, p<0.001). By comparison, LVM/EDV has a substantially numerically weaker correlation (R^2^=0.746, p<0.001, Figure 3).

**Table 1.** Baseline characteristics of the healthy population.

|  | **All** | **Females** | **Males** |
|---|---|---|---|
| Subjects, n (%) | 4461 (100) | 2245 (50) | 2216 (50) |
| Age, years | 59±10 | 59±10 | 59±10 |
| Height, cm | 170±9 | 163±6 | 177±7 |
| Weight, kg | 73±13 | 66±10 | 80±11 |
| BMI, kg/m<sup>2</sup> | 25±3 | 25±4 | 26±3 |
| BSA, m<sup>2</sup> | 1.9±0.2 | 1.7±0.2 | 2.0±0.2 |
| LVM, g | 92±24 | 74±12 | 111±20 |
| LVMI, g/m<sup>2</sup> | 50±10 | 43±6 | 56±8 |
| EDV, mL | 143±32 | 123±21 | 163±29 |
| LVM/EDV, g/mL | 0.65±0.10 | 0.61±0.08 | 0.69±0.10 |
| GT, mm | 6.33±0.90 | 5.71±0.55 | 6.95±0.74 |

Data are presented as mean±SD. Abbreviations: BMI = body mass index; BSA = body surface area; LVM = left ventricular mass; LVMI = left ventricular mass indexed for body surface area; GT = global average left ventricular end-diastolic wall thickness.

**Figure 2.**
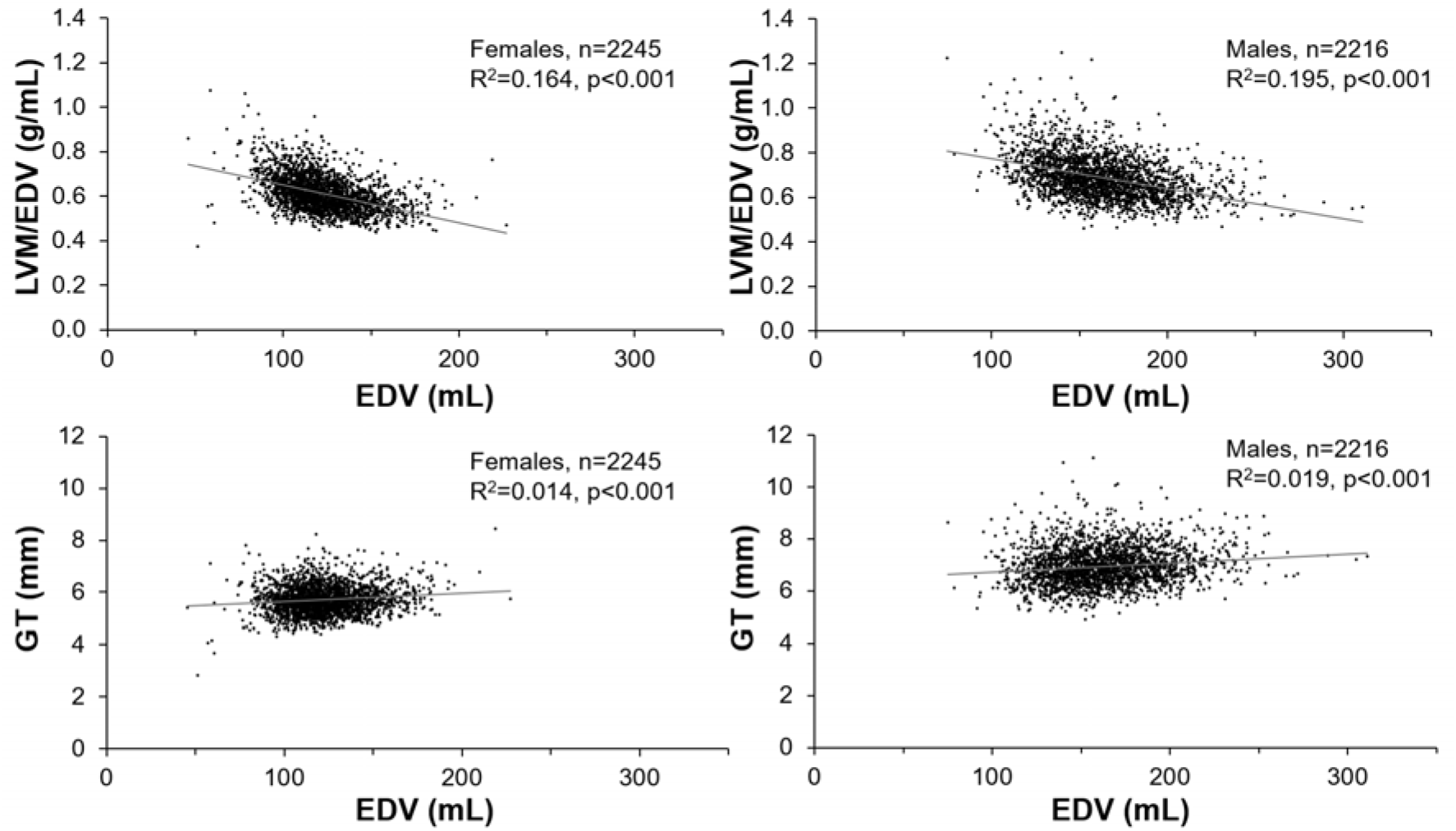
EDV correlated modestly with LVM/EDV but negligibly with GT for healthy subjects of both sexes.

**Figure 3.**
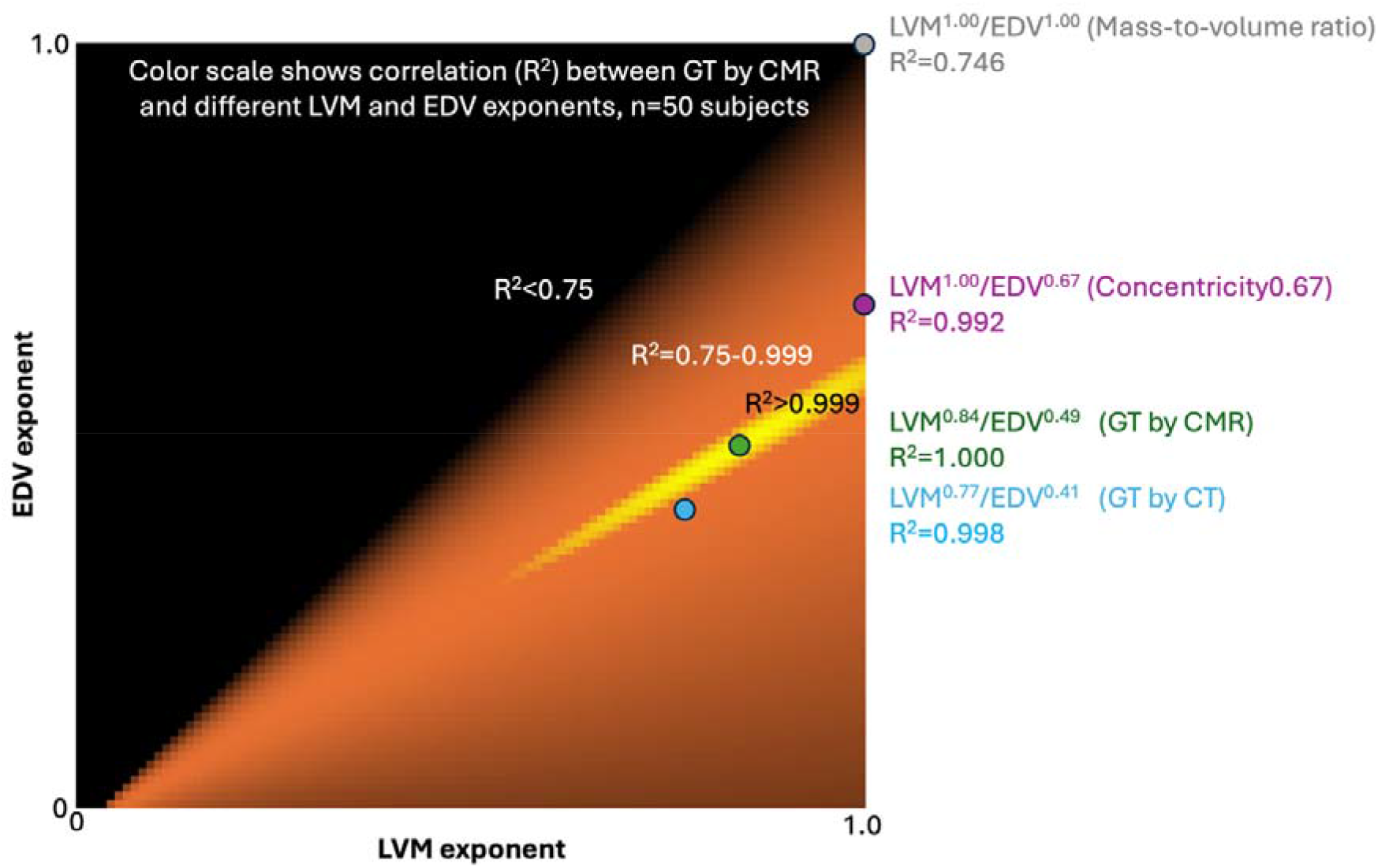
Correlation, expressed as R^2^, between GT by CMR and different exponents of LVM and EDV as depicted by the color scale in a random dataset of 50 healthy subjects undergoing CMR. Black denotes R^2^<0.75, orange denotes R^2^=0.75-0.999, and yellow denotes R^2^>0.999. By definition, GT from CMR (green dot, LVM^0.84^/EDV^0.49^) had an identical correlation (R^2^=1.0). High correlations were found for LVM^0.77^/EDV^0.41^ (blue dot, R^2^=0.992, GT from CT [10]) and LVM/EDV^0.67^ (R^2^=0.992, purple dot, Concentricity^0.67^ [15]. By comparison LVM^1.00^/EDV^1.00^ had a substantially numerically lower correlation (grey dot, R^2^=0.746, mass-to-volume ratio). Note that the yellow region describes the sizable range of combinations of LVM and EDV exponents that all yield an R2>0.999 for the correlation with GT by CMR.

### Intervention populations

#### Changes in LVM and EDV

Baseline characteristics of the interventional study populations stratified by underlying cardiovascular conditions are summarized in Table 2. LVM decreased post-intervention in all four cohorts whereas EDV did not for all groups (see Figure 4). EDV decreased post-intervention only for hypertension, athletes, and aortic regurgitation but it did not differ in aortic stenosis. Figure 5 shows that absolute change in post-intervention LVM and EDV varied across all four cohorts.

**Table 2.** Baseline characteristics of the interventional study populations as stratified by cardiovascular conditions.

|  | **Aortic stenosis** | **Hypertension** | **Athletes** | **Aortic regurgitation** |
|---|---|---|---|---|
| Number, n | 102 | 78 | 28 | 58 |
| Age, years | 69±10 | 58±11 | 27±6 | 56±17 |
| Male, % | 63 (54) | 32 (41) | 23 (82) | 50 (86) |
| Height, cm | 168±9 | 161±9 | 177±8 | 176±9 |
| Weight, kg | 80±18 | 71±18 | 78±14 | 85±17 |
| BMI, kg/m<sup>2</sup> | 28±5 | 27±6 | 25±5 | 27±4 |
| BSA, m<sup>2</sup> | 1.89±0.02 | 1.7±0.2 | 2.0±0.2 | 2.0±0.2 |
| LVM, g | 174±59 | 114±47 | 130±28 | 193±54 |
| LVMI, g/m<sup>2</sup> | 91±26 | 64±19 | 66±10 | 99±25 |
| EDV, mL | 127±50 | 137±40 | 190±32 | 303±76 |
| LVEF, % | 71±16 | 60±9 | 60±5 | 52±9 |
| LVM/EDV, g/mL | 1.41±0.32 | 0.83±0.17 | 0.65±0.11 | 0.64±0.13 |
| GT, mm | 11.4±2.01 | 7.60±1.56 | 7.31±1.02 | 8.13±1.32 |

Data are presented as mean±SD. Abbreviations: BMI = body mass index; BSA = body surface area; LVM = left ventricular mass; LVMI = left ventricular mass indexed for body surface area; EDV = left ventricular end-diastolic volume; ESV = left ventricular end-systolic volume; LVEF = left ventricular ejection fraction; GT = global average left ventricular end-diastolic wall thickness.

**Figure 4.**
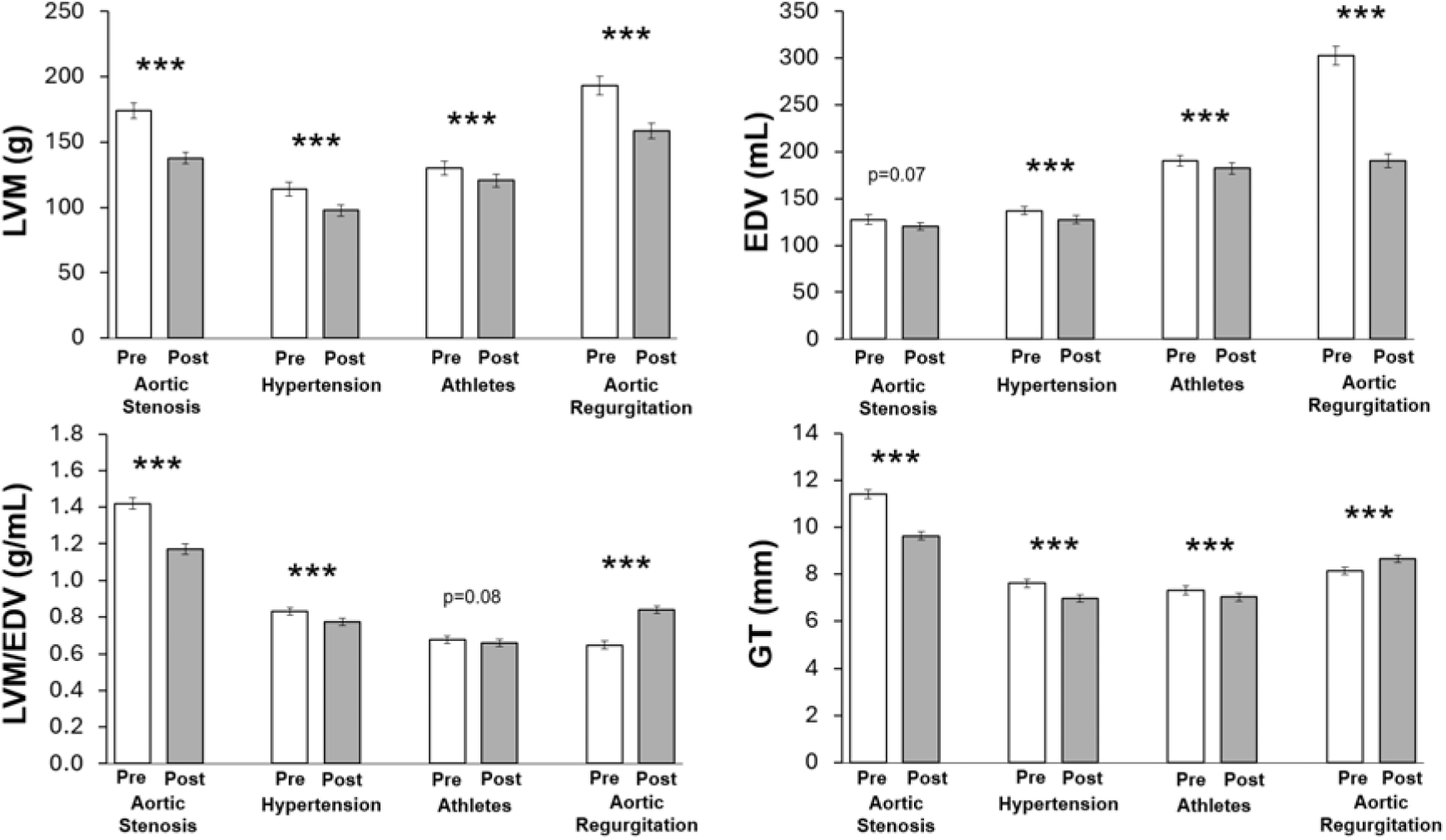
Pre -and post-intervention LVM, EDV, LVM/EDV and GT across all four cohorts. Note LVM and EDV decrease across all four cohorts at different magnitudes, resulting in a numerical decrease in LVM/EDV for aortic stenosis, hypertension and athletes, but an increase in LVM/EDV and GT for the aortic regurgitation group. However, note that change in LVM/EDV in the athlete cohort did not reach statistical significance. Data is presented as mean±SEM.

**Figure 5.**
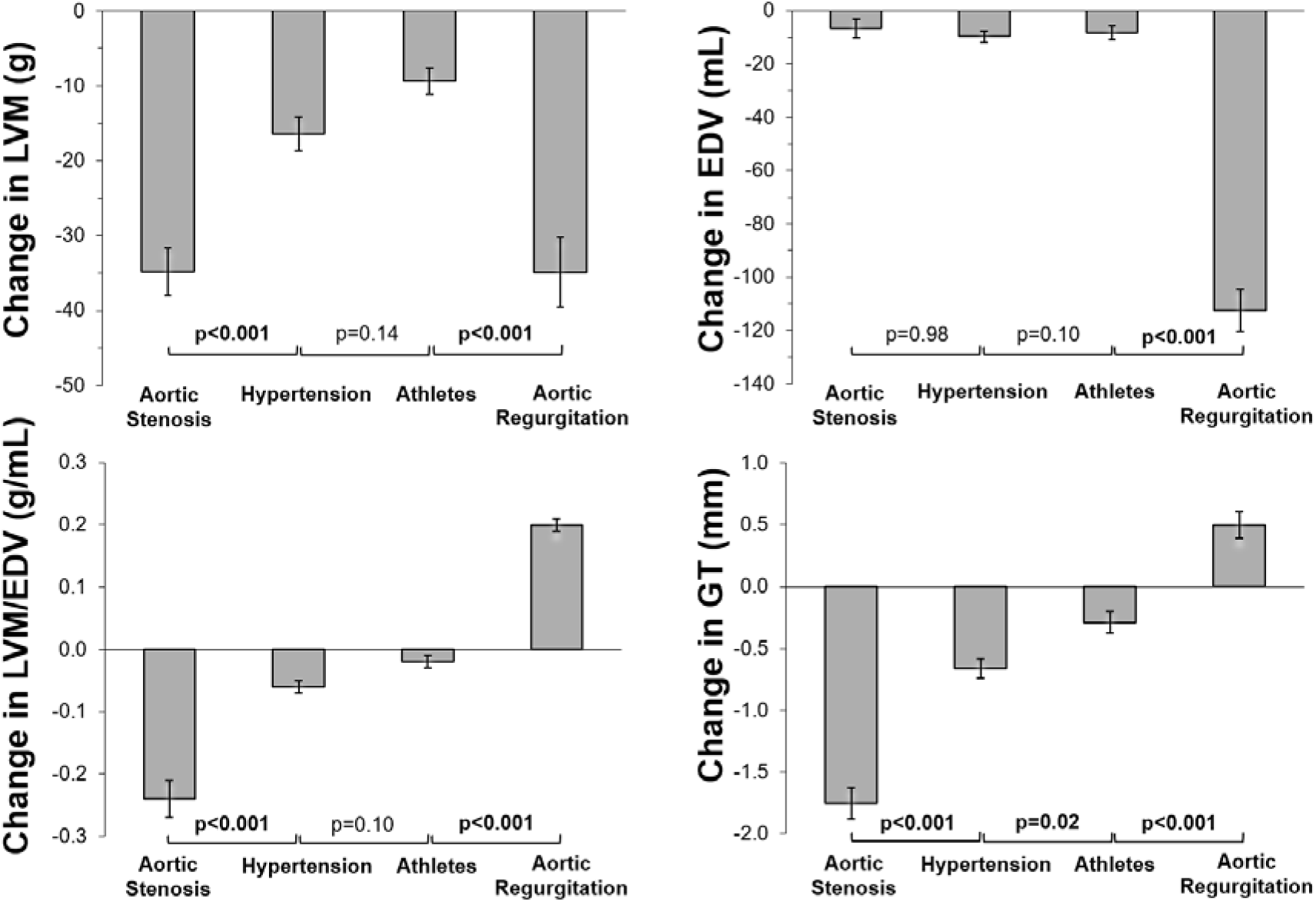
Change in LVM, EDV, LVM/EDV and GT across all four cohorts. Data is presented as mean±SEM. Note that change in GT but not LVM/EDV showed a statistically significant difference across all groups.

#### Changes in LVM/EDV and GT

Both LVM/EDV and GT increased post-intervention for aortic regurgitation but decreased for all other cohorts (Figure 4). GT denotes the directional change in myocardial wall thickness rather than change in overall myocyte mass. GT was able to detect post-intervention changes consistently across all four cohorts, whereas LVM/EDV was not (Figure 4). Also, only GT and not LVM/EDV was able to detect differences in post-intervention change between all four cohorts (Figure 5).

The study on treating hypertension showed that sacubitril/valsartan led to a greater regression in diffuse interstitial fibrosis and LVM by more than two-fold as compared to valsartan alone at one year, but no difference was detected in LVM/EDV between treatment arms. The current analyses showed reduction following intervention for LVM (118±8 vs 97±6g, p<0.001), EDV (138±7 vs 126±5mL, p<0.001), LVM/EDV (0.84±0.02 vs 0.76± 0.02g/mL, p<0.001) and GT (7.76±0.25 vs 6.91±0.21mm, p<0.001). The valsartan group saw reductions in LVM (110±7 vs 99±7g, p<0.001), EDV (136±6 vs 129±7mL, p=0.01), LVM/EDV (0.81±0.03 vs 0.77±0.03g/mL, p=0.004) and GT (7.45±0.25 vs 6.98±0.25mm, p<0.001).

Notably, when comparing absolute changes post-intervention in valsartan alone, sacubitril/valsartan led to a greater reduction in GT (-0.86±0.12 vs -0.46±0.08mm, p=0.007) but the numerical reduction in LVM/EDV did not achieve statistical significance (-0.08±0.02 vs -0.04±0.01g/mL, p=0.06) (Figure 6).

**Figure 6.**
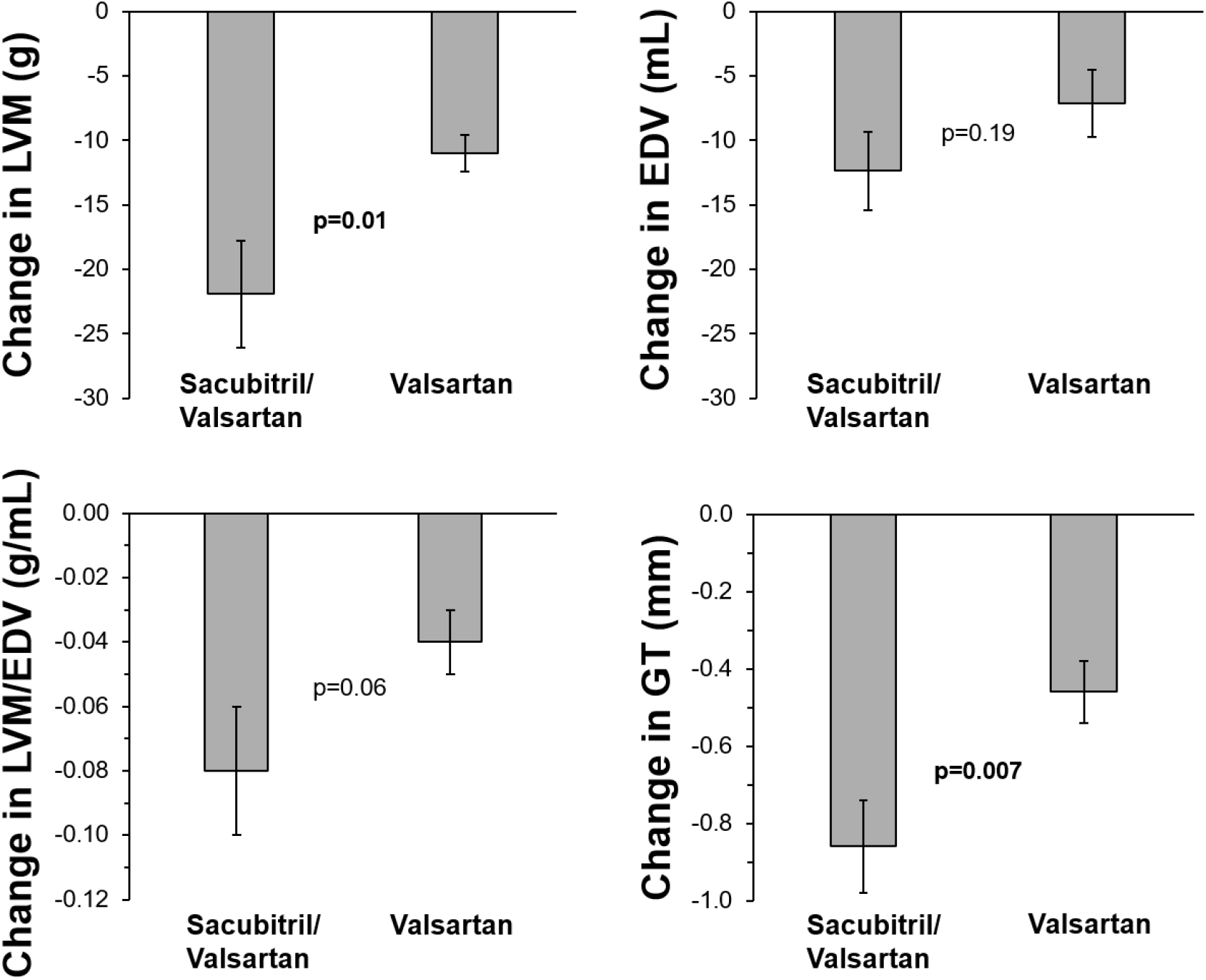
Change in LVM, EDV, LVM/EDV and GT in sub-analysis of the hypertensive cohort as stratified by treatment arms (sacubitril/valsartan versus valsartan alone). There is a reduction post-intervention in all measures, however, only LVM and GT reach statistical significance to detect a change between treatment arms. Data is presented as mean±SEM.

## Discussion

The main findings of this study were that GT in health remains effectively unchanged across the range of EDV. This indicates that constancy of average LV wall thickness (GT) is inherent to healthy human physiology, thus making GT an appropriate measure for evaluating pathophysiology impacting the balance between left ventricular volume and mass. Furthermore, GT is more sensitive than LVM/EDV at detecting changes following intervention across four physiologically distinct conditions ranging from aortic stenosis, untreated hypertension, athletes, and aortic regurgitation.

### GT in health

Normal reference values for GT among 4461 healthy subjects in the current study were 4.6–6.8 mm and 5.5–8.4 mm for females and males, respectively. These findings are highly consistent with previous findings among 99 healthy subjects of 4.8–7.1 mm and 5.8–8.5 mm for females and males, respectively [8], and the small differences are likely due to the differences in sample size, population, and the improved precision related to fully automated artificial intelligence-based delineation in the current study.

### Estimates of the balance between LVM and EDV

Changes in LVM are difficult to interpret without taking into account EDV. Therefore, in assessing LVH, one needs to take into account the balance of LVM and EDV, and how both change. There are a number of CMR measures that have attempted to consider the balance between LVM and EDV. Firstly, GT takes into account the non-linear relationship between LVM, EDV and wall thickness (Figure 1), and GT has previously been validated to be a highly accurate and precise estimate of average LV wall thickness [8]. Secondly, LVM/EDV is a commonly reported measure of the balance between LVM and EDV but incorrectly assumes a linear relationship between LVM and EDV. An elevated LVM/EDV is commonly used to describe concentric remodelling. This is based on the simplistic reasoning that an increase in LV mass that is relatively greater than the increase in LV volume results in a higher LVM to EDV ratio. However, an increase in EDV that is greater than the increase in LVM can lead to a lower LVM/EDV, which reflects eccentric remodelling. Thirdly, LVM/EDV^2/3^, also referred to as concentricity^0.67^, is also used to describe LVH. LVM/EDV^2/3^ was developed in 2010 to refine the definition of LVH by considering concentricity of the left ventricle. LVM/EDV^2/3^ is theoretically preferable over LVM/EDV as it scales mass and volume to the same geometric dimension, taking into account that wall volume of a sphere grows at a square rate unlike volume of a sphere which grows at a cubic rate [15]. However, LVM/EDV^2/3^ reflects an uncalibrated geometric relationship between LVM and EDV with unintuitive units (g/ml^0.67^), whereas GT provides a calibrated measure of absolute average LV wall thickness in millimetres. This arguably positions GT as a more intuitive measure for use in everyday clinical practice. Also, the prognostic value of GT has been shown to be higher than for LVM/EDV^2/3^ in LVH [8]. GT has the added unique advantage over LVM/EDV of reflecting directional change, where a positive change in GT denotes an increase in average LV wall thickness and vice versa.

### Change in LV hypertrophy in aortic stenosis

The current study shows that after AVR in aortic stenosis, there was a reduction in LVM, LVM/EDV and GT, but not EDV. Previous studies have shown that AVR reduces LVM, EDV, and LVM/EDV in aortic stenosis [17]. However, the current findings show that AVR in aortic stenosis leads to by far the greatest reduction in GT compared to the other studied LV hypertrophy interventions (Figure 4).

### Change in LV hypertrophy in hypertension

The current study showed that LVM, EDV, LVM/EDV and GT decreased after antihypertensive treatment. LVM regression is achievable with antihypertensive treatments [18], and it has been shown to be able to reverse concentric remodelling, albeit as measured by LVM/EDV [19]. The current study is the first to demonstrate that GT reduces with treatment of hypertension. In order to determine the true net effect on change in LVH, we argue that beyond change in LVM, concurrent change in EDV must also be considered. In considering both LVM and EDV, we showed that only GT was able to detect a difference between treatment arms in this study, whereas LVM/EDV did not. This finding adds empirical evidence to the theoretical advantages of GT, also suggesting that GT provides improved precision over LVM/EDV in estimating changes in either wall thickness as the balance between LVM and EDV.

### Change in LV hypertrophy in athletes

The current study has shown a reduction in both LVM and EDV in athletes undergoing forced detraining. LVM regression in detraining has been consistently demonstrated [13] [20], whereas EDV reduction has not [20]. The current study found an ability to detect a reduction in hypertrophy with GT and not LVM/EDV following one month of forced detraining. While not many studies have examined the change in LVM/EDV post-detraining, one longitudinal study with a median follow-up of 12 years did show a reduction in LVM/EDV [21]. The small magnitude of the change in GT (0.3 mm [4%] reduction) supports that notion that athletic remodelling largely preserves wall thickness (GT), and we hypothesise that monitoring of GT gives the most correct assessment of cardiac remodelling. Another prospective longitudinal study using echocardiography observed that over a 90-day period, competitive endurance and strength training resulted in different effects on the cardiac structure. They showed that both endurance and strength training resulted in increased LVM of similar magnitude, but only endurance training resulted in concomitant increase in EDV. Therefore, accounting for an increase in EDV, endurance athletes did not experience an increase in LV wall thickness while strength athletes did [22]. However, it has been suggested that training type (endurance vs strength) alone does not reliably predict cardiac remodelling based on currently accepted CMR measures [23].

### Change in LV hypertrophy in aortic regurgitation

The current study has shown that aortic valve replacement reduced both LVM and EDV in aortic regurgitation, and this is consistent with previous studies [24] [25]. However, despite the decrease in both LVM and EDV, the current study also shows that GT *increased* post-intervention, and GT is arguably the only measure that accurately reflects the change in wall thickness on balance between the changes in LVM and EDV. The study from which the current GT calculations have been made is the only study that has used CMR to study the change in the left ventricle following aortic valve replacement for aortic regurgitation [14]. While there are a number of echocardiographic studies looking at left ventricular changes post-AVR, there was only one small study (n=8) that reported no change in interventricular septal wall thickness measured by echocardiography [26]. The differences between that study and the current study emphasises the differences in precision between linear measurement of wall thickness by echocardiography and estimation of GT using three-dimensional volumetric measurements of LVM and EDV by CMR.

### Comparing changes in GT between conditions

Cardiac remodelling is a chronic adaptive response to biomechanical stresses and is influenced by neurohumoral factors and intracellular signalling cascades. This ultimately leads to changes in myocardial composition and structure. Cardiomyocyte adaptation to biomechanical stress is intended to preserve an ideal fibre or wall sheer stress [27]. The cardiomyocyte is able to thicken or elongate by adding new sarcomeres either in parallel or in series, respectively [28]. Haemodynamic simulations have established that increased fibre stress causes an increase in wall thickness while increased wall shear stress causes dilatation [27]. As expected, both increased thickness and length of the cardiomyocyte would lead to an increase in LVM and EDV, but at different magnitudes. This geometric change eventually leads to concentric or eccentric LVH. The pathophysiology that underlies specific cardiovascular conditions results in varying degrees of fibre versus wall shear stress. This consequently shapes cardiac remodelling and LV geometry. This is clearly depicted in valvular heart disease, whereby increase in afterload (aortic stenosis) leads to increased wall thickness, while increase in volume (aortic regurgitation) leads to predominantly increased EDV. In both conditions, cardiomyocytes add new sarcomeres therefore increasing LVM. The key difference lies within the structure of remodelling, whether sarcomeres are added in parallel or in series, which in turn affects the magnitude of change in EDV. Expectedly, these two fundamentally differing conditions induce contrasting net effects on wall thickness measured as GT despite the increase in both LVM and EDV. In a less extreme example, normal physiological adaptations in athletes involve different degrees of change to LVM and EDV depending on the nature of exercise, which is not necessarily pathological. Taken together, the interplay between fibre and wall shear stress shapes the ratio of change between LVM and EDV, and which on balance are uniquely accurately reflected in GT.

### GT as a better measure of true change in LVH

Cardiomyocyte adaptation to biomechanical stress is intended to preserve an ideal myocardial fibre stress or wall shear stress [27]. We hypothesize that this may explain the findings of GT as a physiological constant in health, since it reflects an ideal target wall thickness in normal cardiac physiology. Our results suggest that an intraindividual change in GT during the development of pathological processes may represent an underlying pathophysiological adaptation which predates an increase in LVM or EDV above the upper limits of normal. This is further supported by previous findings that GT is the LV hypertrophy metric with the strongest association with prognosis among patients with otherwise normal range LVMI and EDVI [8]. Indeed, low normal EDVI combined with high normal LVMI values result in GT values above the normal range. In assessing for the change in LV hypertrophy, the current findings indicate that LVM should not be monitored in isolation as every change in LVM concomitantly results in some degree of change in EDV. Therefore, change in LVM alone does not represent the true net effect of change in hypertrophy. Importantly, in comparing LVM/EDV and GT, GT was the only measure able to detect a change in all four cardiovascular conditions and in between treatment arms for hypertension. Finally, figure 3 illustrates that there are several different combinations of exponents for LVM and EDV that all yield an exceptionally strong and close correlation with the CMR GT formula used in the current study, but LVM/EDV had a substantially lower correlation, illustrating the underlying differences between the measures.

## Limitations

This was a retrospective study of pre-existing CMR measures with a mixture of manual delineations and automated artificial intelligence-based delineations from different CMR labs. This is reasonable as this is the first study exploring the role of GT in change in LVH post-intervention. Future prospective studies using GT would provide more robust clinical evidence. This study compared various CMR indices. Studies on relative strength of association of these and outcomes are warranted.

## Conclusions

GT, unlike LVM/EDV, is a precise estimate of average LV wall thickness that is effectively physiologically constant in health and can be used to track pathological changes in hypertrophy. GT is reliable and reproducible in detecting post-intervention change in a variety of cardiovascular conditions, taking into account the balance between LVM and EDV. Instead of LVM/EDV, GT should preferentially be used to quantify changes in LV hypertrophy over time.

## Clinical Perspectives

CMR is considered the non-invasive reference standard for accurate and reproducible quantification of LV volumes and mass, and we introduce the previously validated measure of wall thickness, GT, for evaluating net effects of changes in LVM and EDV following interventions. The advantage of GT lies in detecting subtle changes to LVH by taking into account the balance of changes in both LVM and EDV, thereby having a distinct role in detecting early changes in LVH. The current study suggests that an intraindividual change in GT reflects a pathological process with better accuracy than LVM/EDV.

## Data Availability

All data produced in the present study are available upon reasonable request to the authors

## Acknowledgements

The authors acknowledge and thank co-authors of the five published studies [9] [11] [12] [13] [14] that comprise patient populations reanalysed and presented in this study. The authors also acknowledge and thank the PRECISION-HCM Collaborative for collating data from the UK Biobank, Framingham Heart Study Offspring Cohort, MyoFit46 and several other independent studies based in London, United Kingdom, from which the subgroup of healthy normal participants were selected for this study.

## Abbreviations List

AVR: aortic valve replacement
CMR: cardiovascular magnetic resonance
EDV: left ventricular end-diastolic volume
GT: global average left ventricular end-diastolic wall thickness
LVH: left ventricular hypertrophy
LVM: left ventricular mass

